# Sex-Related Differences in Long-term Outcomes across the Spectrum of Ejection Fraction in Heart Failure Patients

**DOI:** 10.1101/2023.09.26.23296192

**Authors:** Akane Kawai, Yuji Nagatomo, Midori Yukino-Iwashita, Yukinori Ikegami, Makoto Takei, Ayumi Goda, Takashi Kohno, Atsushi Mizuno, Mitsunobu Kitamura, Shintaro Nakano, Munehisa Sakamoto, Yasuyuki Shiraishi, Shun Kohsaka, Takeshi Adachi, Tsutomu Yoshikawa, WET-HF investigators

## Abstract

**Background:** Recently, patients with supra-normal left ventricular ejection fraction (snLVEF) are reported to have high risk of adverse outcomes, especially in women. We sought to evaluate sex-related differences in the association between LVEF and long-term outcomes in heart failure (HF) patients.

**Methods:** The multicenter WET-HF Registry enrolled all patients hospitalized for acute decompensated HF (ADHF). We analyzed 3,943 patients (age 77 years; 59.8% male) registered from 2006 to 2017. According to LVEF the patients were divided into the 3 groups: HF with reduced EF (HFrEF), mildly reduced EF (HFmrEF) and preserved EF (HFpEF). The primary endpoint was defined as the composite of cardiac death and ADHF rehospitalization after discharge.

**Results:** Implementation of guideline-directed medical therapy (GDMT) such as renin-angiotensin-system inhibitor (RASi), β-blocker and their combination at discharge was significantly lower in women than men in HFmrEF. Lower prescription of RASi + β-blocker combination in female HFmrEF was noted even after adjustment for covariates (p=0.007). There were no such sex-related differences in HFrEF. Female sex was associated with higher incidence of the primary endpoint and ADHF rehospitalization after adjustment for covariates exclusively in HFmrEF. Restricted cubic spline analysis demonstrated a U-shaped relationship between LVEF and the hazard ratio of the primary endpoint in women, but such relationship was not observed in men (*p* for interaction=0.037).

**Conclusions:** In women, not only lower LVEF but also snLVEF were associated with worse long-term outcomes. Additionally, sex-related differences in the GDMT implementation for HFmrEF highlight the need for sex-specific guidelines to optimize HF management.

## Introduction

Heart failure (HF) has been increasing in prevalence and is a leading cause of death; in addition, HF poses a considerable socioeconomic burden worldwide. Even with the development of novel agents and devices for the management of HF, clinical outcomes have not sufficiently improved over a decade.^1^

Substantial differences are known to exist in the characteristics and clinical course of HF between male and female patients; these differences are important but underrecognized.^2^ Female patients with HF are older and have a lower body mass index (BMI), less ischemic heart disease,^3^ and poorer quality of life.^4^ Moreover, guideline-directed medical therapy (GDMT) is less likely to be implemented in female patients with HF and reduced ejection fraction (HFrEF), which is related to their older age, worse renal function, and lower body weight compared with male patients.^5^ In contrast, women are more likely to have HF with preserved EF (HFpEF) ^6, 7^ and higher left ventricular ejection fraction (LVEF) compared with men.^8–10^ In epidemiological data from Europe, the prevalence of HFpEF was higher in women than in men and increased with age.^11^ More recently, the proportion of HFpEF was shown to increase over time, with women outnumbering men approximately 2:1 among HF cases between 2000 and 2010 in the Olmsted County, Minnesota, USA.^12^ A higher prevalence of HFpEF in women than in men has been consistently reported in Japan.^3, 7^

Recently, supra-normal LVEF (snLVEF) has been identified as a population associated with a potentially high risk of future cardiovascular adverse events. In the general population, snLVEF is associated with higher all-cause mortality compared with the LVEF 60%-65% group in both sexes, but this trend was more evident in women.^13^ On the other hand, compared with male patients, female patients with snLVEF who underwent coronary computed tomography had worse outcomes.^14^ However, long-term outcomes of snLVEF in patients with HF have not yet been fully investigated.

Therefore, in the present study, we aimed to evaluate sex-related differences in the association between LVEF and long-term outcomes in patients with HF in a prospective multicenter registry. We also aimed to evaluate sex differences in the implementation of GDMT in patients with HFrEF that potentially affect sex disparities in clinical outcomes.

## Methods

### Study Design and Sample Population

This is the retrospective observational study from Japanese multicenter registry. The details of the West Tokyo Heart Failure Registry (WET-HF) registry have been previously described. ^15^ This database is a prospective, multicenter cohort registry designed for the collection of data pertaining to the clinical backgrounds and outcomes of patients hospitalized with acute decompensation heart failure (ADHF) who fulfilled the Framingham criteria. ^16^

The seven participating institutes are located in Tokyo and Saitama, including three university hospitals (Keio University, Kyorin University, and Saitama Medical University) and four tertiary referral hospitals (Sakakibara Heart Institute, St. Luke’s International Hospital, Saiseikai Central Hospital, and National Hospital Organization Tokyo Medical Center). ADHF was diagnosed by individual cardiologists at each institution according to the Framingham criteria ^16^; patients with acute coronary syndrome or isolated right-sided HF were excluded from the registry. Baseline data and outcomes for the WET-HF Registry were collected by dedicated clinical research coordinators from medical records and interviews with treating physicians to obtain a robust assessment of patient care and patient outcomes. Data were entered into an electronic data-capturing system with a robust data query engine and system validations for data quality. Outliers in continuous variables or unexpected values in categorical variables were identified based on established criteria, and the originating institution was contacted to verify the values. The quality of reporting was also verified by the principal investigators (Y.S. and S.K.) at least once a year, and periodic queries were conducted to ensure quality. Exclusive on-site auditing by the principal investigators ensured proper registration of each patient.^17–19^

Before the launch of the WET-HF Registry, information regarding the objective of the present study, its social significance, and an abstract were provided for clinical trial registration with the University Hospital Medical Information Network (UMIN000001171). The study protocol was approved by the institutional review boards at each site, and research was conducted in accordance with the Declaration of Helsinki. Ethical approval for the study was granted by the institutional review boards of Keio University Hospital (#20090176) and all participating institutions. Written and/or oral informed consent was obtained from each patient before registration. Information on medications at admission and discharge were prospectively collected. GDMT was defined as prescriptions of renin-angiotensin-system inhibitors (RASi), β-blockers, and mineralocorticoid receptor antagonists (MRA) because novel agents such as angiotensin receptor neprilysin inhibitor (ARNI) and sodium-glucose cotransporter 2 (SGLT-2) inhibitors were not available in Japan during this study period. Echocardiography was performed during the index hospitalization after stabilization of HF signs and symptoms. LVEF was evaluated by modified Simpson method at each institution. LV end-diastolic volume and end-systolic volume were calculated using the Teichholz method ^20^ and indexed by body surface area.

We analyzed 4,000 patients who were enrolled in the WET-HF Registry from January 2006 to December 2017. Out of 4,000 patients, 57 patients were excluded because of a lack of echocardiogram data (**Figure 1**). As a result, 3,943 patients were included for analysis in the present study. The patients were divided into the three groups according to LVEF: HFrEF (LVEF < 40%; n = 1604), HFmrEF (40≤ LVEF <50%; n = 707) and HFpEF (LVEF ≥ 50%; n = 1,632). The proportion of female patients was 29% in the HFrEF group, 39% in the HFmrEF group and 52% in the HFpEF group (**Figure 1**).

**Figure 1.**
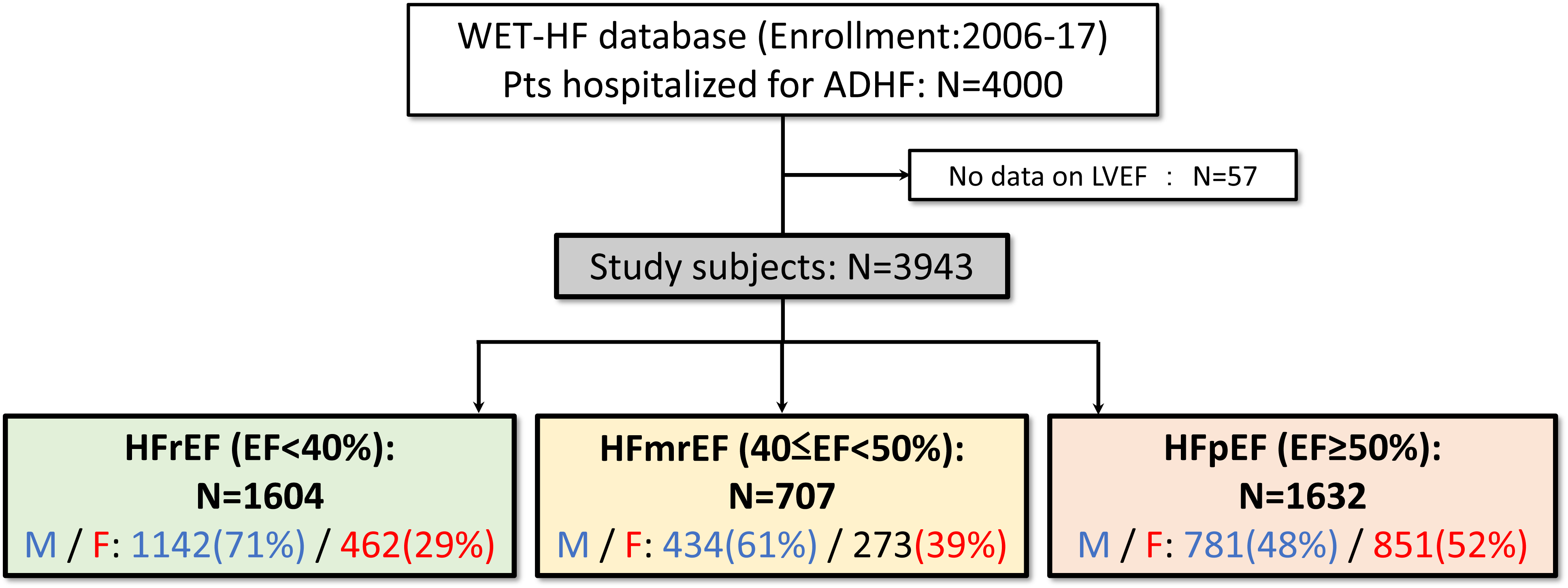
Flowchart of the Study. WET-HF, West Tokyo Heart Failure Registry; ADHF, acute decompensated heart failure; M, male; F, female; HFrEF, heart failure with reduced ejection fraction; HFmrEF, heart failure with mildly reduced ejection fraction; HFpEF, heart failure with preserved ejection fraction; LVEF, left ventricular ejection fraction.

### Endpoints

A follow-up survey using medical charts or telephone interviews was performed, and patients who were lost to follow-up were censored at the date of last contact. Regarding HF readmission, treating physicians at each participating hospital made decisions according to the usual standard of care. The date of index hospitalization discharge, ADHF rehospitalization, and death were properly collected and confirmed by site investigators and dedicated clinical research coordinators. The WET-HF registry is supported by a central study committee that adjudicates mode of death. Initially, all deaths were reviewed by the investigators and then categorized into those in need of adjudication and those whose mode of death could be defined clearly. Central committee members (S.K., Y.S., T.K., Y.N., A.G., and T.Y.) reviewed the abstracted record and adjudicated the mode of death.^18^ For the present study, the primary endpoint of long-term outcome was defined as a composite of cardiac death and ADHF rehospitalization during 1,000 days after discharge.

### Statistical Analysis

Continuous variables were expressed as mean ± standard deviation for normally distributed data and as median (interquartile range) for data with non-normal distribution. Between-group differences were assessed using t-test or Wilcoxon test for unpaired data, as appropriate. Multivariable logistic regression analysis was performed to determine whether sex was associated with the implementation of GDMT by adjusting for the covariates such as age, history of ADHF admission, etiology of HF (DCM/ischemic/VHD), systolic blood pressure (SBP) at discharge, estimated glomerular filtration rate (eGFR) at admission, and LVEF. Multivariate Cox proportional hazards model analysis was performed to determine the independent predictor of the primary endpoint adjusting for age, BMI, etiology of HF, concomitant atrial fibrillation (AF), systolic blood pressure at admission, eGFR at admission, hemoglobin level (Hb), B-type natriuretic peptide (BNP) at admission, LVEF, and geriatric nutritional risk index (GNRI). The restricted cubic spline curves show the function relating LVEF to the primary endpoint, where LVEF 60% is set to the reference value (hazard ratio [HR] = 1). These analyses were also performed after exclusion of patients with VHD, since LVEF could be affected by specific hemodynamics in VHD (e.g. overestimation of LVEF in mitral regurgitation). ^21^

The aforementioned analysis was carried out using R version 4.2.1 (R Foundation for Statistical Computing, Vienna, Austria) with the “rms” package. Other statistical analyses were performed using JMP 15.2.0 (SAS Institute Inc., Cary, NC, USA). Statistical significance was set at *p* < 0.05.

## Results

Baseline characteristics comparing male and female patients in the three groups of HFrEF, HFmrEF and HFpEF are shown in **Table 1**. In each group, female patients were older and had lower BMI, lesser prevalence of ischemic etiology, lower Hb level, and smaller LV chamber size. **Supplemental Table 1** shows in-hospital treatment, which was almost similar between men and women. **Table 2** shows in-hospital outcome, and vital signs, laboratory data and medication at discharge. In-hospital outcome was also similar between both sexes. **Figure 2A** shows the prescription rate of GDMT at discharge in the HFrEF and HFmrEF. The prescription rate of RASi, β-blockers and their combination was lower in women than in men in the HFmrEF group, whereas there were no significant differences in the HFrEF group (**Figure 2A** and **Table 2**). Both in HFrEF and HFmrEF patients, the dose of β-blockers (*p* < 0.001) were lower in women than in men (HFrEF: male vs. female, 5.0 mg vs. 2.5 mg, p <0.001; HFmrEF: 3.8 mg vs. 2.5 mg, p <0.001, **Figure 2B** and **Table 2**). Multivariable logistic regression analysis demonstrated that female sex was associated with lower prescription of RASi plus β-blockers in the HFmrEF group (**Figure 2D** and **Table 2**), whereas there was no significant difference in GDMT prescription between both sexes in the HFrEF group (**Figure 2C** and **Table 2**).

**Table 1.** Baseline characteristics of the study population comparing men and women in HFrEF, HFmrEF and HFpEF.

|  | HFrEF<br>N = 1,604 |  |  | HFmrEF<br>N = 707 |  |  | HFpEF<br>N = 1,632 |  |  |
| --- | --- | --- | --- | --- | --- | --- | --- | --- | --- |
|  | Male<br>(n = 1142) | Female<br>(n = 462) | *p*-value | Male<br>(n = 434) | Female<br>(n = 273) | *p*-value | Male<br>(n = 781) | Female<br>(n = 851) | *p*-value |
| Demographics |  |  |  |  |  |  |  |  |  |
| Age, years | 71 (60-80) | 79 (69-85) | <0.001 | 75 (65-83) | 80 (72-86) | <0.001 | 78 (70-85) | 82 (75-87) | <0.001 |
| BMI, kg/m<sup>2</sup> | 21.8 (19.6-24.4) | 19.9 (17.7-22.5) | <0.001 | 21.5 (19.4-24.1) | 20.4 (17.9-22.9) | <0.001 | 22.0 (19.7-24.2) | 20.7 (18.0-23.5) | <0.001 |
| Etiology of heart failure |  |  |  |  |  |  |  |  |  |
| Ischemic, n (%) | 458 (40) | 142 (31) | <0.001 | 198 (46) | 78 (29) | <0.001 | 185 (24) | 89 (10) | <0.001 |
| DCM, n (%) | 361 (32) | 105 (23) | <0.001 | 43 (10) | 16 (6) | 0.053 | 12 (1.5) | 9 (1.1) | 0.39 |
| Valvular, n (%) | 106 (9) | 92 (20) | <0.001 | 72 (17) | 94 (34) | <0.001 | 283 (36) | 391 (46) | <0.001 |
| History and comorbidities |  |  |  |  |  |  |  |  |  |
| History of HF admission,<br>n (%) | 391 (35) | 163 (36) | 0.67 | 103 (24) | 83 (30) | 0.065 | 202 (26) | 238 (28) | 0.31 |
| HT, n (%) | 727 (64) | 288 (62) | 0.62 | 310 (72) | 189 (69) | 0.50 | 540 (69) | 550 (65) | 0.053 |
| DM, n (%) | 458 (40) | 157 (34) | 0.024 | 173 (40) | 75 (27) | <0.001 | 258 (33) | 232 (27) | 0.011 |
| AF, n (%) | 472 (41) | 161 (35) | 0.017 | 184 (43) | 131 (48) | 0.16 | 425 (54) | 485 (57) | 0.28 |
| Stroke or TIA, n (%) | 178 (16) | 54 (12) | 0.044 | 51 (12) | 29 (11) | 0.65 | 113 (14) | 116 (14) | 0.67 |
| COPD, n (%) | 61 (5) | 10 (2) | <0.01 | 29 (7) | 2 (1) | <0.01 | 69 (9) | 25 (3) | <0.01 |
| SAS, n (%) | 24 (17) | 10 (13) | 0.52 | 10 (19) | 4 (9) | 0.38 | 8 (9) | 7 (6) | 0.63 |
| Pacemaker Implantation, n (%) | 66 (6) | 41 (9) | 0.028 | 33 (8) | 34 (12) | 0.035 | 55 (7) | 66 (8) | 0.58 |
| ICD implantation, n (%) | 84 (7) | 22 (5) | 0.052 | 11 (3) | 5 (2) | 0.53 | 10 (1) | 9 (1) | 0.68 |
| CRT Implantation, n (%) | 39 (3) | 15 (3) | 0.86 | 4 (0.9) | 1 (0.4) | 0.37 | 1 (0.1) | 1 (0.1) | 0.95 |
| Vital signs |  |  |  |  |  |  |  |  |  |
| sBP, mmHg | 132 (111-154) | 131 (110-153) | 0.38 | 140 (123-164) | 134 (120-160) | 0.013 | 140 (120-165) | 138 (120-162) | 0.33 |
| Heart rate, beats/min | 96 (78-115) | 96 (78-116) | 0.77 | 92 (76-111) | 96 (77-120) | 0.18 | 80 (67-101) | 88 (72-110) | <0.001 |
| Laboratory data |  |  |  |  |  |  |  |  |  |
| eGFR, mL/min/1.73m<sup>2</sup> | 48.6 (33.2-62.8) | 48.3 (32.9-64.6) | 0.62 | 48.7 (34.4-62.6) | 48.7 (33.1-64.6) | 0.81 | 51.1 (32.8-65.2) | 49.4 (33.8-66.0) | 0.66 |
| Hb, g/dL | 13.2 (11.5-14.9) | 11.9 (10.4-13.3) | <0.001 | 12.4 (10.7-14.0) | 11.3 (9.8-12.7) | <0.001 | 11.6 (10.0-13.3) | 11.1 (9.8-12.7) | <0.001 |
| HbA1c, % | 6.0 (5.6-6.6) | 6.0 (5.5-6.6) | 0.57 | 6.0 (5.5-6.7) | 5.8 (5.5-6.3) | 0.012 | 5.8 (5.4-6.4) | 5.7 (5.4-6.2) | 0.002 |
| Na, mEq/L | 140 (137-142) | 140 (137-142) | 0.41 | 140 (138-142) | 140 (137-143) | 0.56 | 140 (137-142) | 140 (137-142) | 0.89 |
| K, mEq/L | 4.4 (4.0-4.8) | 4.3 (3.9-4.6) | <0.001 | 4.3 (4.0-4.7) | 4.3 (3.9-4.7) | 0.19 | 4.3 (3.9-4.7) | 4.2 (3.9-4.6) | 0.031 |
| BNP, pg/μL | 885 (485-1406) | 1041 (578-1834) | 0.001 | 650 (354-1256) | 859 (412-1556) | 0.012 | 492 (275-905) | 509 (275-912) | 0.84 |
| NT-proBNP, pg/μL | 5633<br>(3057-11952) | 6873<br>(3743-15086) | 0.043 | 3995<br>(2073-8700) | 5563<br>(2869-13033) | 0.016 | 2289<br>(1121-4073) | 2960<br>(1449-5963) | <0.001 |
| BNP/NT-proBNP quartile, n (%) | 162/248/340/357<br>(15/22/31/32) | 40/87/136/183<br>(9/20/30/41) | <0.001 | 106/112/104/98<br>(25/27/25/23) | 43/63/66/91<br>(16/24/25/35) | 0.003 | 306/214/123/101<br>(41/29/17/14) | 296/228/181/115<br>(36/28/22/14) | 0.030 |
| GNRI | 100 (92-107) | 94 (86-102) | <0.001 | 97 (89-105) | 95 (88-103) | 0.056 | 99 (92-105) | 95 (87-103) | <0.001 |
| Echocardiography |  |  |  |  |  |  |  |  |  |
| LVEF, % | 29 (23-34) | 31 (25-35) | <0.001 | 45 (42-46) | 45 (42-46) | 0.95 | 60 (55-63) | 60 (56-64) | 0.016 |
| LVDd, mm | 60 (55-66) | 55 (48-60) | <0.001 | 53 (48-58) | 49 (44-53) | <0.001 | 48 (43-54) | 44 (39-48) | <0.001 |
| LVDs, mm | 52 (46-58) | 46 (40-52) | <0.001 | 40 (35-45) | 37 (32-41) | <0.001 | 32 (28-37) | 29 (25-32) | <0.001 |
| LAD, mm | 45 (40-50) | 42 (37-47) | <0.001 | 43 (39-48) | 43 (38-48) | 0.48 | 45 (40-50) | 44 (39-49) | 0.031 |
| E/e' | 16.9 (12.0-23.9) | 18.1 (11.0-26.0) | 0.35 | 14.4 (10.7-20.9) | 16.8 (10.0-28.0) | 0.038 | 15.2 (10.4-21.0) | 19.0 (12.1-27.7) | <0.001 |
| TRPG, mmHg | 29 (21-37) | 27 (21-36) | 0.25 | 26 (21-35) | 30 (24-37) | 0.003 | 29 (23-39) | 32 (25-41) | 0.006 |

HF<sub>r</sub>EF, heart failure with reduced ejection fraction; HF<sub>m</sub>rEF, heart failure with mildly reduced ejection fraction; HF<sub>p</sub>EF, heart failure with preserved ejection fraction; BMI, body mass index; sBP, systolic blood pressure; DCM, dilated cardiomyopathy; HT, hypertension; DM, diabetes mellitus; AF, atrial fibrillation; TIA, transient ischemic attack; ICD, implantable cardioverter defibrillator; CRT, cardiac resynchronization therapy; eGFR, estimated glomerular filtration rate; Hb, hemoglobin; Na, serum sodium level; K, serum potassium level; B-type natriuretic peptide; NT-proBNP, N-terminal pro-brain natriuretic peptide; GNRI, Geriatric Nutritional Risk Index; LVEF, left ventricular ejection fraction; LVDd, left ventricular end-diastolic diameter; LVDs, left ventricular end-systolic diameter; LAD, left atrial diameter; TRPG, tricuspid regurgitant pressure gradient.

**Table 2.** In-hospital outcomes and vital signs, laboratory data and medication at discharge.

|  | HFrEF<br>N = 1,604 |  |  | HFmrEF<br>N = 707 |  |  | HFpEF<br>N = 1,632 |  |  |
| --- | --- | --- | --- | --- | --- | --- | --- | --- | --- |
|  | Male<br>(n = 1142) | Female<br>(n = 462) | *p*-value | Male<br>(n = 434) | Female<br>(n = 273) | *p*-value | Male<br>(n = 781) | Female<br>(n = 851) | *p*-value |
| In-hospital outcomes |  |  |  |  |  |  |  |  |  |
| Hospital stay (days) | 16 (11-26) | 16 (11-26) | 0.90 | 14 (10-23) | 15 (10-24) | 0.60 | 13 (9-22) | 14 (9-22) | 0.43 |
| In-hospital death | 46 (4) | 23 (5) | 0.40 | 13 (3) | 11 (4) | 0.46 | 21 (3) | 30 (4) | 0.33 |
| Vital signs and laboratory data at discharge |  |  |  |  |  |  |  |  |  |
| sBP, mmHg | 107 (96-118) | 107 (96-120) | 0.59 | 114<br>(102-128) | 100<br>(100-122) | 0.021 | 116<br>(104-128) | 112 (102-124) | <0.001 |
| Heart rate, bpm | 71 (62-80) | 72 (64-81) | 0.049 | 70 (61-78) | 70 (64-82) | 0.043 | 69 (60-77) | 70 (62-78) | 0.002 |
| eGFR, | 50.7<br>(34.5-64.5) | 49.1<br>(34.3-65.1) | 0.78 | 48.8<br>(34.8-62.3) | 49.3<br>(32.1-64.9) | 0.652 | 51.2<br>(34.6-64.9) | 49.2 (34.2-64.0) | 0.39 |
| Hb, g/dl | 12.9<br>(11.2-14.7) | 11.7<br>(10.6-13.1) | <0.001 | 12.3<br>(10.5-13.9) | 11.3<br>(10.0-12.7) | <0.001 | 11.6<br>(10.4-13.1) | 11.2 (10.0-12.6) | <0.001 |
| K, mEq/L | 4.4 (4.1-4.6) | 4.3 (4.0-4.6) | 0.018 | 4.4 (4.1-4.7) | 4.3 (3.9-4.6) | 0.007 | 4.3 (4.0-4.6) | 4.3 (3.9-4.6) | 0.08 |
| Medication at discharge |  |  |  |  |  |  |  |  |  |
| Loop diuretic, n (%) | 850 (78) | 357 (82) | 0.096 | 309 (73) | 181 (69) | 0.23 | 552 (73) | 604 (74) | 0.65 |
| Dose, mg | 20 (20-40) | 20 (20-40) | 0.056 | 20 (20-40) | 20 (20-40) | 0.76 | 20 (20-40) | 20 (20-40) | 0.021 |
| RASi, n (%) | 751 (69) | 301 (69) | 0.99 | 279 (66) | 154 (59) | 0.049 | 458 (60) | 461 (56) | 0.10 |
| ACEi, n (%) | 425 (57) | 160 (53) | 0.29 | 115 (41) | 63 (41) | 0.93 | 170 (37) | 159 (34) | 0.41 |
| ARB, n (%) | 335 (45) | 142 (47) | 0.45 | 166 (60) | 95 (62) | 0.66 | 295 (64) | 306 (66) | 0.53 |
| β-B, n (%) | 948 (87) | 379 (86) | 0.87 | 348 (83) | 196 (75) | 0.014 | 474 (62) | 550 (67) | 0.050 |
| Carvedilol/Bisoprolol/Other, n (%) | 715/227/6<br>(75/24/1) | 276/100/3<br>(73/26/1) | 0.60 | 227/111/10<br>(65/32/3) | 113/80/3<br>(58/41/2) | 0.084 | 248/208/18<br>(52/44/4) | 225/307/18<br>(41/56/3) | 0.001 |
| β-B dose, mg carvedilol | 5.0 (2.5-7.5) | 2.5 (1.3-5.0) | <0.001 | 3.8 (2.5-5.0) | 2.5 (1.3-5.0) | <0.001 | 2.5 (2.5-5.0) | 2.5 (1.9-5.0) | 0.011 |
| MRA, n (%) | 485 (44) | 202 (46) | 0.54 | 125 (30) | 93 (36) | 0.12 | 191 (25) | 222 (27) | 0.38 |
| Tolvaptan, n (%) | 68 (7) | 27 (7) | 0.82 | 5 (1) | 12 (5) | 0.010 | 36 (5) | 40 (5) | 0.88 |
| GDMT |  |  |  |  |  |  |  |  |  |
| RASi + βB + MRA, n (%) | 340 (31) | 131 (30) | 0.64 | 76 (18) | 46 (18) | 0.87 | 90 (12) | 88 (11) | 0.49 |
| RASi + βB, n (%) | 682 (62) | 271 (62) | 0.82 | 238 (57) | 114 (44) | <0.001 | 299 (39) | 322 (39) | 0.98 |

HFREF, heart failure with reduced ejection fraction; HFmrEF, heart failure with mildly reduced ejection fraction; HFpEF, heart failure with preserved ejection fraction; PDE-III, phosphodiesterase III; sBP, systolic blood pressure; DCM, dilated cardiomyopathy; eGFR, estimated glomerular filtration rate; Hb, hemoglobin; K, serum potassium level;
RASi, renin-angiotensin-system inhibitor; ACE, angiotensin converting enzyme inhibitor; ARB, angiotensin receptor antagonist; β-B, β-blocker; MRA, mineralocorticoid receptor antagonist; GDMT, guideline-directed medical therapy.

**Figure 2.**
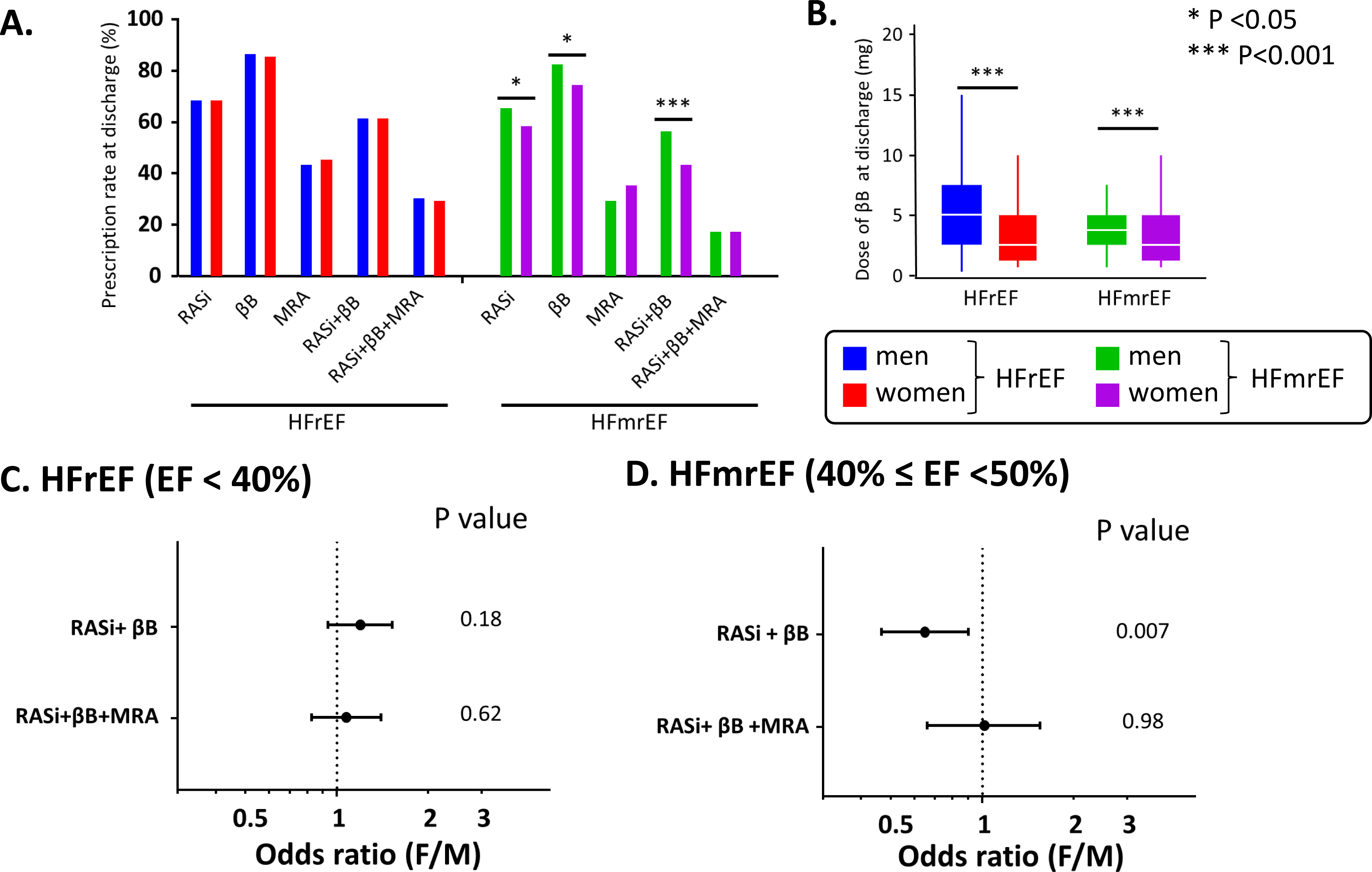
The Prescription of GDMT at Discharge in Men and Women. **A.** The prescription rate of each medication and their combination. **B**. Dose of β-blocker at discharge (mg carvedilol). **C** and **D**. Multivariable logistic regression analysis depicting association of sex with GDMT implementation in the patients with HFrEF (**C**) and HFmrEF (**D**). RASi, renin-angiotensin system inhibitor; βB, β-blocker; MRA, mineralocorticoid receptor antagonist; HFrEF, heart failure with reduced ejection fraction; HFmrEF, heart failure with mildly reduced ejection fraction; GDMT, guideline-directed medical therapy; LVEF, left ventricular ejection fraction

**Supplemental Figure** shows the Kaplan-Meier curves for the primary endpoint (cardiac death and ADHF rehospitalization) according to sex in the whole population (**Supplemental Figures A, B,** and **C**), in the HFrEF (**Supplemental Figures D, E,** and **F**), in the HFmrEF (**Supplemental Figures G, H,** and **I**), and in the HFpEF (**Figures 3J, K,** and **L**) subgroups. Female patients had a higher incidence of the primary endpoint and ADHF rehospitalization than male patients in the whole population (**Supplemental Figures A and C**). Such difference was also observed in the HFmrEF group, but it showed borderline significance in the primary endpoint (**Supplemental Figures G** and **I**). The sex difference was not statistically significant in the HFrEF group (**Supplemental Figures D** and **F)**. The incidence of cardiac death was similar between sexes in the whole population and all subgroups (**Supplemental Figures B, E, H** and **K**). Multivariable Cox proportional hazard model analysis showed that sex did not remain an independent predictor of the primary endpoint (female/male, HR 1.09, 95% CI: 0.91–1.31, *p* = 0.34, **Figure 3A**) or ADHF rehospitalization (HR 1.13, 95% CI: 0.94–1.37, *p* = 0.19, **Figure 3B**) after adjustment for the covariates. However, in the HFmrEF group, female sex was independently associated with higher incidence of the primary endpoint (HR 1.62, 95% CI: 1.05–2.50, *p* = 0.029, **Figure 3A**) and ADHF rehospitalization (HR 1.76, 95% CI: 1.12–2.76, *p* = 0.014, **Figure 3B**). There was no significant difference in incidence of these endpoints between both sexes in HFrEF or HFpEF (**Figures 3A** and **B**).

**Figure 3.**
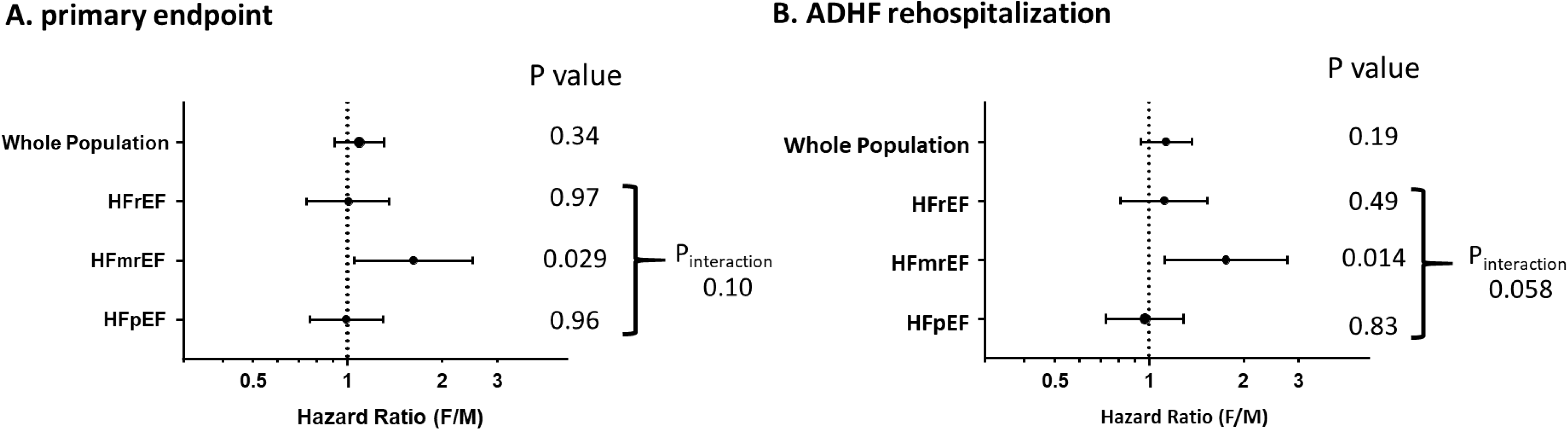
Cox Proportional Hazard Model Analysis Evaluating Hazard Ratio (F/M) of the Primary Endpoint (A) and ADHF Rehospitalization (B) between Both Sexes in the Whole Population, HFrEF, HFmrEF and HFpEF Subgroups. ADHF, acute decompensated heart failure; F, female; M, male; HFrEF, heart failure with reduced ejection fraction; HFmrEF, heart failure with mildly reduced ejection fraction; HFpEF, heart failure with preserved ejection fraction.

### Relationship between LVEF and Hazard Ratio of the Primary Endpoint

We evaluated hazard ratios of the primary endpoint among groups divided by 10 percentage units of LVEF, in female and male patients. In female patients, the hazard ratio was higher in the LVEF < 20% group and 70% ≤ LVEF group (LVEF < 20%: HR, 2.63; 95% CI: 0.96–6.05; 70% ≤ LVEF: HR, 1.83; 95% CI: 0.99–3.20; **Figure 4C**), but it did not reach significance. After exclusion of valvular heart disease (VHD), the hazard ratios of the LVEF < 20% and 70% ≤ LVEF groups were significant in female patients (LVEF < 20%: HR, 3.59; 95% CI: 1.24–9.06, p=0.021; 70% ≤ LVEF: HR, 2.19; 95% CI: 1.01–4.44; p=0.047, **Figure 4F**). However, these associations were not observed in male patients (**Figure 4B** and **E**). We found a significant interaction between sex and LVEF groups (*p* for interaction = 0.013 for the whole group; *p* = 0.039 for the non-VHD group).

**Figure 4.**
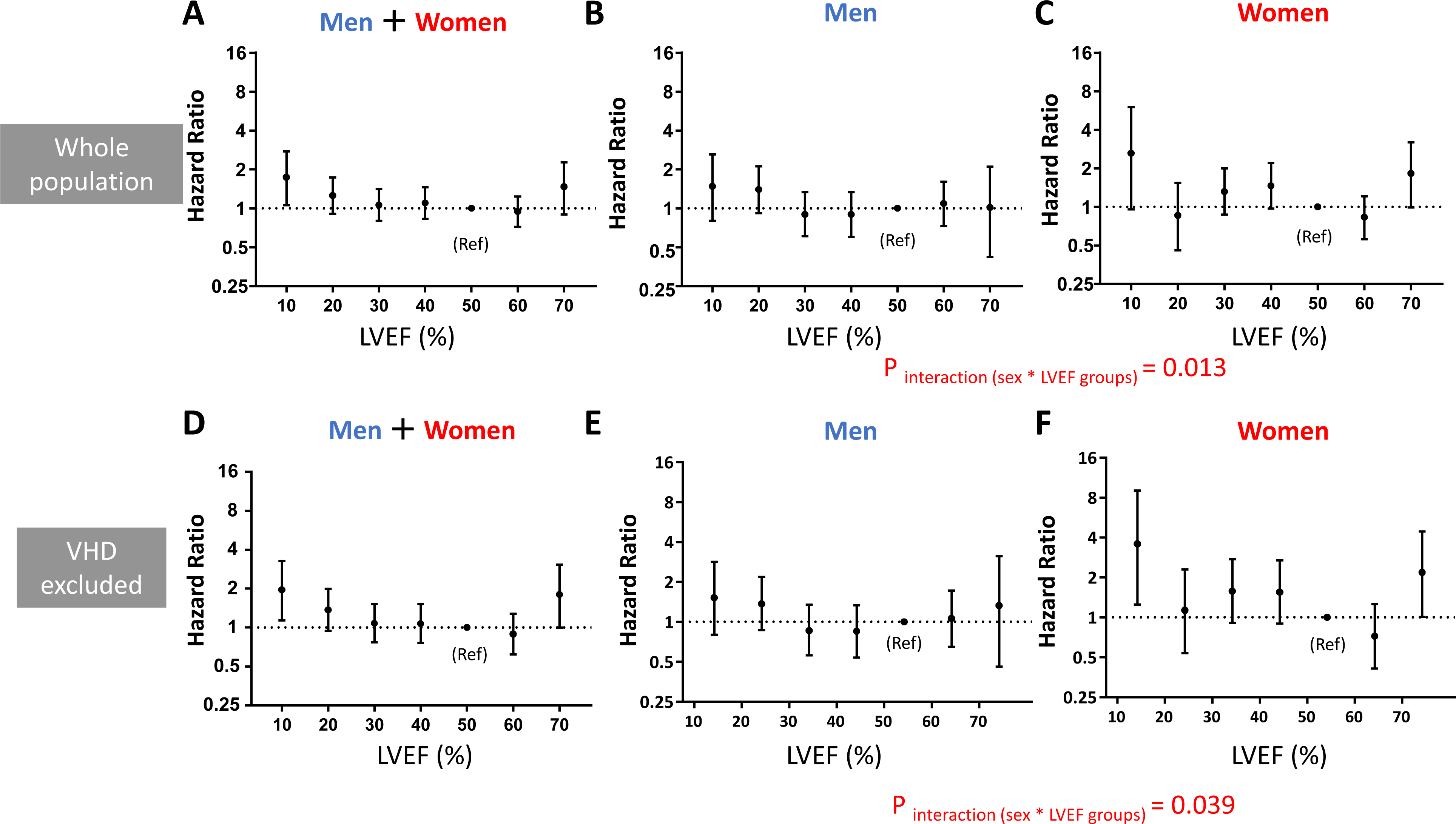
The Relationship between LVEF and Hazard Ratio of the Primary Endpoint in Both Sexes (A, D), Men (B, E), and Women (C, F) in Whole Population (Upper Panel) and Those with Non-valvular Etiology (Lower Panel). The patients were divided into groups divided by 10 percentage units of LVEF. The reference group was 50% ≤ LVEF < 60%. LVEF, left ventricular ejection fraction; Ref, reference.

### Restricted Cubic Spline Curve for the Relationship between LVEF and Hazard Ratio of the Primary Endpoint

The relationship between LVEF and the hazard ratio of the primary endpoint was evaluated using restricted cubic spline analysis. In female patients, it showed a U-shaped relationship with a nadir at LVEF 60%, whereas such a trend was not observed in male patients (**Figures 5B** and **C**). A significant interaction between sex and LVEF was observed (*p* for interaction = 0.037). After exclusion of VHD, the U-shaped relationship in female patients was more evident, and the hazard ratio for lower and higher LVEF groups was significant. Such a relationship was not observed in male patients (sex × LVEF interaction: *p* = 0.012; **Figures 5E** and **F**).

**Figure 5.**
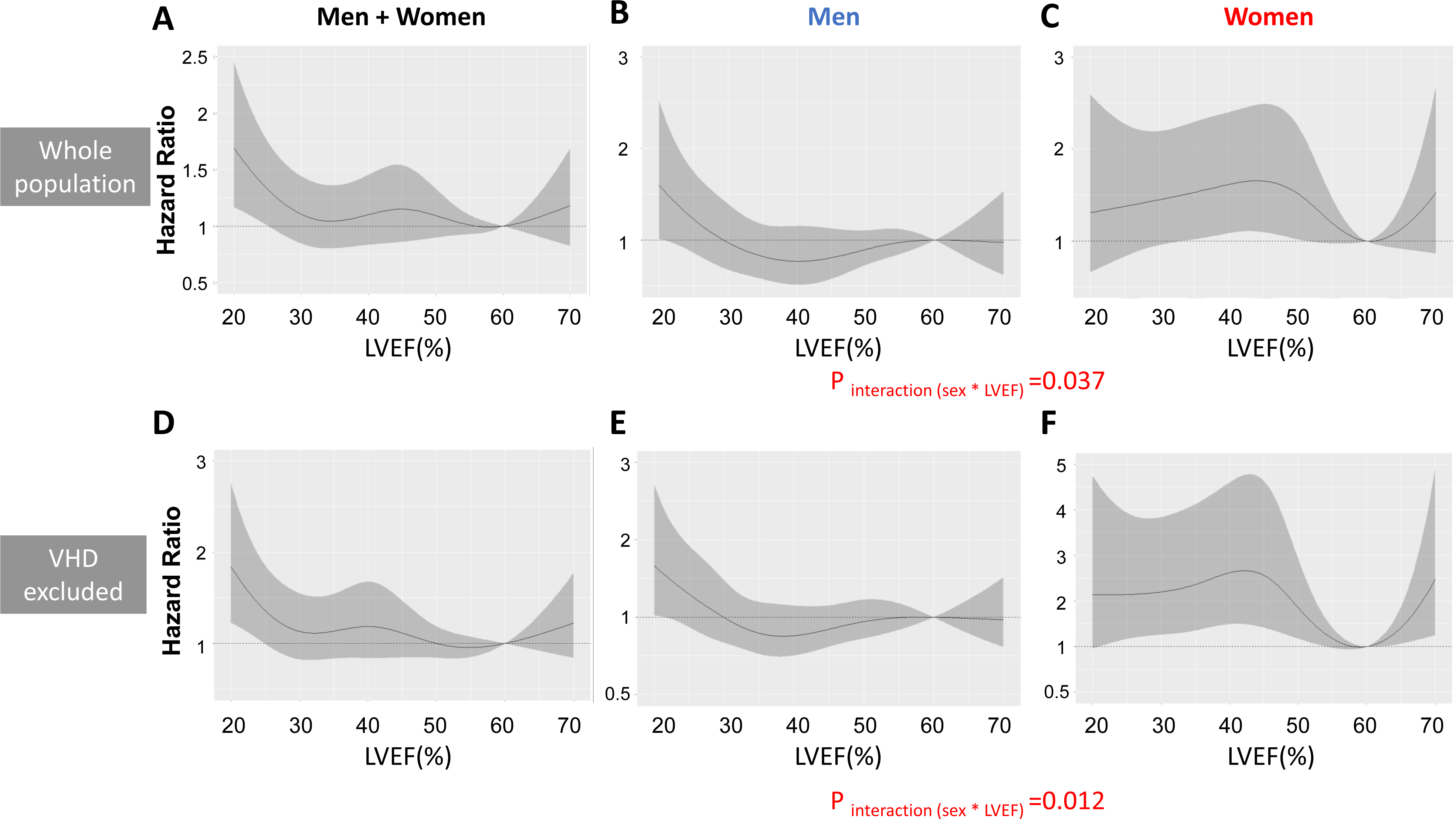
Restricted Cubic Spline Analysis for the Relationship between LVEF and the Primary Endpoint in Both Sexes (A, D), Men (B, E), and Women (C, F) in Whole Population (Upper Panel) and Those with Non-valvular Etiology (Lower Panel). The restricted cubic spline curve depicting the relationship between LVEF and hazard ratios of primary endpoint showed a U-shaped relationship between LVEF and hazard ratio of the primary endpoint in female patients (**C**) but not in male patients (**B**). After exclusion of valvular heart disease, the U-shaped relationship became more evident, and the hazard ratios of lower and higher LVEF were significant in female patients (**F**), but not in male patients (E). LVEF, left ventricular ejection fraction; VHD, valvular heart disease.

### Characteristics in HFpEF Further Divided by LVEF in Male and Female Patients

The baseline characteristics in HFpEF further divided by LVEF group are shown in **Supplemental Table 2**. There were no significant differences in most variables, including demographics, comorbidities, and laboratory data, among 3 LVEF groups in female and male patients, except for AF and B-type natriuretic peptide/N-terminal pro-brain natriuretic peptide (BNP/NT-pBNP). In both sexes, LV chamber size characteristics such as LVDd and LV end-diastolic volume index (LVEDVi) were smaller in higher LVEF groups. Across all LVEF groups, LVDd and LVEDVi were consistently smaller in female patients than in male patients (**Supplemental Table 2**).

## Discussion

In the present study, we demonstrated the following main findings. First, female sex was associated with a lower rate of GDMT implementation in HFmrEF. Second, female patients had a higher incidence of the primary endpoint and ADHF rehospitalization during long-term follow-up in the HFmrEF after adjusting for covariates. Third, the association of LVEF with hazard ratio of the primary endpoint showed a U-shaped relationship in women. This trend was not observed in men and a significant interaction between sexes and LVEF was observed. Especially, after exclusion of VHD, the elevated hazard ratio in lower and higher LVEF was significant in women. Finally, whereas LV chamber size was smaller in higher LVEF groups, it was consistently smaller in women than in men across all LVEF groups.

### Sex-related Difference in the Association of lower LVEF with Clinical Outcome

In the present study, even mildly lower LVEF was associated with higher incidence of the primary endpoint in women, whereas such trend was not observed in men. Of note, in the present study, a lower GDMT implementation in women was seen in HFmrEF rather than HFrEF (**Figure 2**). It was consistently observed even after adjustment for confounding factors (**Figure 2D**). The potential mechanism of that observed exclusively in female HFmrEF remains unknown, although lower implementation of GDMT in female HFrEF has been reported in the previous studies. ^5^. Since the efficacy of GDMT on HFmrEF patients has been controversial and HFmrEF might be perceived by clinicians as a ‘less severe’ form of HFrEF, the implementation of GDMT might be more likely to be biased by miscellaneous factors such as socioeconomic characteristics ^22^ or a higher likelihood of adverse effects of GDMT in women than in men.^23–25^ The efficacy of GDMT on HFmrEF patients has not been established since no randomized controlled trials (RCTs) were conducted exclusively on this population. However, observational studies, ^26–28^ post-hoc analyses of RCTs, ^29, 30^ and meta-analyses of RCTs ^31, 32^ suggest its potential benefit.

Taken together, these results suggest that the lower prescription rates and insufficient up-titration of GDMT might be related to worse outcomes in patients with even mildly lower LVEF in women (**Figure 6A**).

**Figure 6.**
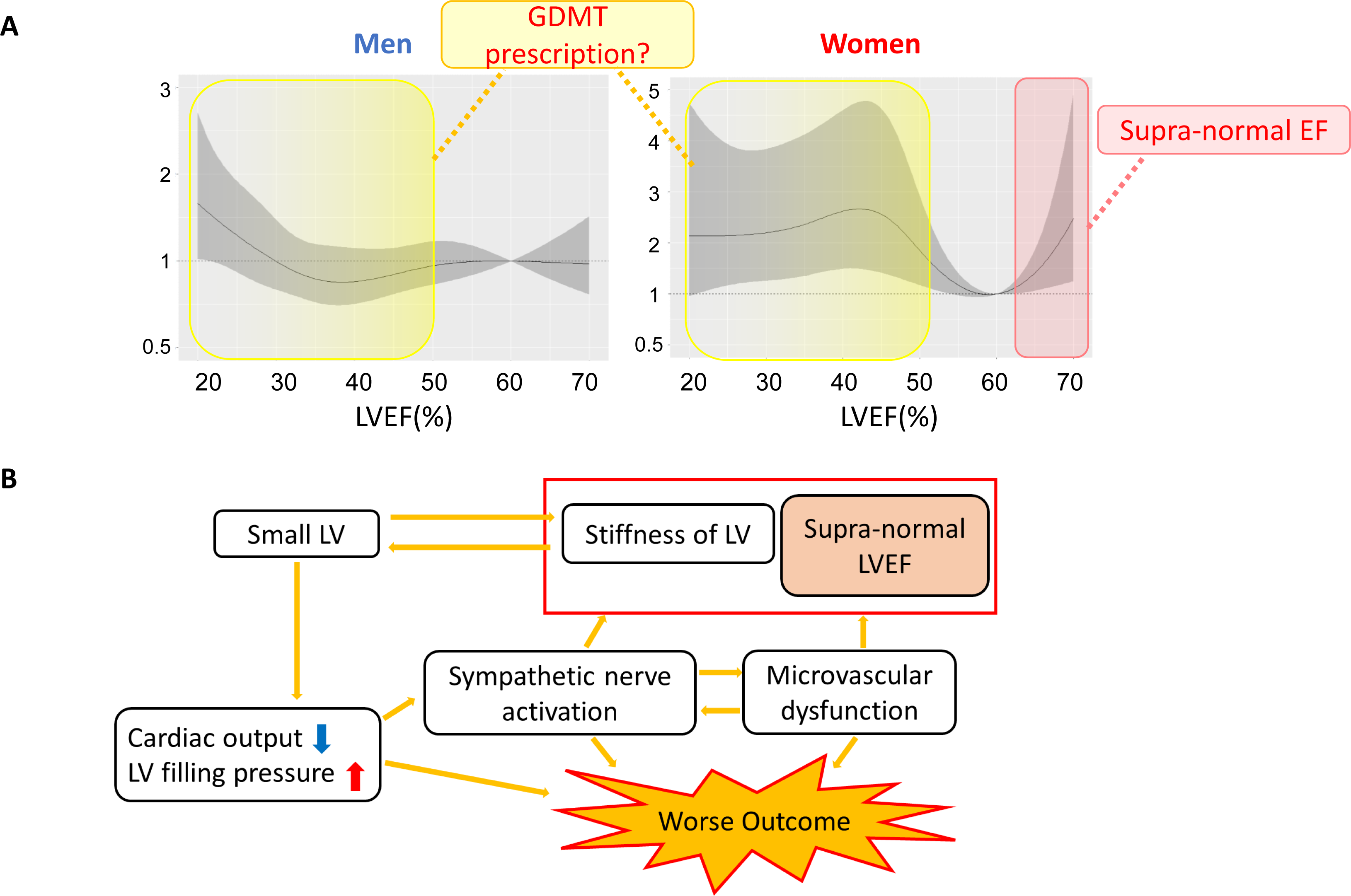
Graphical Summary. In female patients, both lower and higher LVEF were associated with higher hazard ratio of the primary endpoint. Potential mechanisms might include lower prescribing of GDMT for mildly lower LVEF and the pathophysiology of snLVEF for higher LVEF. The snLVEF might reflect LV stiffening rather than enhanced systolic function, as described in the Discussion. Small LV size was also shown to be associated with LV stiffness. Small LV size can cause lower cardiac output and elevated LV filling pressure, which can lead to sympathetic nerve activation. Because microvascular dysfunction and sympathetic nerve activation are associated with snLVEF, especially in women, they might underlie these findings and could explain the worse outcomes of female patients with snLVEF. GDMT, guideline-directed medical therapy; LVEF, left ventricular ejection fraction; snLVEF, supra-normal left ventricular ejection fraction.

### LV Stiffening and Smaller Chamber Size as a Key Feature of snLVEF

Increased LVEF is shown to be associated with higher LV end-diastolic elastance (Eed) as well as end-systolic elastance (Ees), but not with higher preload recruitable stroke work (PRSW), loading condition independent measures of contractility.^9, 33^ These findings indicate that an increase in Ees might be reflective of passive stiffening rather than enhanced systolic function (**Figure 6B**).^33^ In the general population, Eed and Ees increase with age, and the increase is reported to be greater in women than in men.^33^ Further, the positive association of higher Ees or snLVEF with LV stiffness rather than with PRSW has also been confirmed in patients with HFpEF in recent studies that conducted comprehensive echocardiography and invasive exercise testing.^9, 10^ As a result, the changes of end-diastolic pressure-volume relationship (EDPVR) during exercise differs substantially between patients with snLVEF and those with lower LVEF (e.g., LVEF 50% to 60% ^10^ or 65% ^9^). Whereas patients with snLVEF showed a higher increase in Ees and blunted augmentation in LVEDVi which results in left-shifted EDPVR, those with lower LVEF showed right-shifted EDPVR through LV volume expansion, which is an adaptation process more typical of HFrEF.^9, 10^

An increased LVEF is associated with smaller LV chamber size,^9, 10^ which is more prevalent in older women,^34, 35^ as shown in the present study. A smaller LV volume was shown to be associated with higher Ees, indicating a possible association of smaller LV volume with progressing stiffness (**Figure 6B**).^36^ Smaller LV size has been shown to be associated with impaired exercise capacity and cardiac reserve, represented by smaller augmentation in LV stroke volume and cardiac output in healthy women.^37^ Augmentation of LV chamber size during exercise is blunted in female patients with HFpEF compared with male patients, and it results in more elevated LV filling pressure.^38^ From these findings, women with a smaller LV chamber might need to keep hyperdynamic conditions to compensate for its disadvantage. Smaller LV size per se was also associated with higher mortality risk, even in patients with the same LVEF.^13^

### Sex Differences in the Association of snLVEF with Clinical Outcome

In critically ill patients hospitalized in intensive care units, snLVEF (>70%) was reported to be associated with increased adjusted 28-day mortality compared with patients with normal EF.^39^ Recently, a large regional healthcare system based study reported that adjusted hazard ratios for mortality showed a U-shaped relationship for LVEF, with a nadir of risk at an LVEF of 60% to 65%. Although this relationship was observed in both sexes in all age-stratified groups, it was more evident in female patients, with a significant interaction between sexes.^13^ In another study enrolling subjects with EF ≥ 57%, higher LVEF was significantly associated with an increased risk of major adverse cardiovascular events (MACE) among individuals with low but not high stroke volume.^40^ An association between snLVEF and increased mortality might be particularly relevant in the female population, whereas men did not show the same relationship. Other studies enrolling patients who underwent non-invasive imaging modality testing indicate that women with snLVEF had a higher risk of mortality ^14, 41^ and MACE,^42^ whereas men did not show the same relationship. In the present study, we observed a higher incidence of the primary endpoint in female patients with higher LVEF (**Figure 5F** and **6A**), a finding consistent with these previous studies.

The potential mechanism mediating snLVEF and increased mortality in female patients remains unknown. However, according to a study that enrolled patients who underwent ^13^N-ammonia positron emission tomography, women with snLVEF showed reduced coronary flow reserve and blunted heart rate response to adenosine infusion, indicating microvascular dysfunction and heightened sympathetic nerve activity. This association was not observed in men.^42^ As LV hypercontractility and cardiac sympathetic hyperactivity have been observed in patients with coronary microvascular dysfunction,^43^ these features could reflect the mechanism underlying the poorer prognosis in this population (**Figure 6B**). Endothelial inflammation and coronary microvascular dysfunction may be the common pathophysiology among HF syndromes in women,^2^ which may explain the sex-related differences in HF. Sex hormones have been considered as potential candidates mediating this pathophysiology. A previous study suggested that estrogen affects the autonomic nervous system, attenuating sympathetic nervous tone in humans.^44^ Estrogen can also regulate coronary microcirculation by the facilitation of endothelial functions through production of endothelial nitric oxide synthase.^45, 46^ Moreover, a lack of testosterone might cause impaired myocardial perfusion, which has been demonstrated in mice.^47^ Therefore, sex hormones might have meaningful effects on the autonomic nervous system or microvascular dysfunction, and these phenomena could result in worse outcomes in snLVEF only in female patients with snLVEF compared with male patients.

Taken together, worsened LV stiffness, smaller LV volume, microvascular dysfunction, and sympathetic nerve overactivation might, at least in part, account for the increase in cardiac events observed in women with snLVEF (**Figure 6B**).

### Limitations

Our study has several limitations. First, this study was a retrospective study based on observational registry data. Second, LVEF was measured at each institute, which potentially introduces the risk of inter-observer variability. Third, novel agents for treatment of HF (e.g., ARNI, SGLT-2 inhibitors) were not available during this study period, which differs from the current clinical practice. Finally, WET-HF is a registry of the Japanese population, and caution is warranted when applying the present findings to other ethnicities or regions.

## Conclusions

In female patients, not only lower LVEF but also snLVEF were associated with worse long-term outcomes. Further investigations are warranted to elucidate the potential mechanisms underlying poorer outcomes in the female snLVEF population.

Additionally, differences in the implementation of GDMT for HFmrEF patients between men and women highlight the need for sex-specific guidelines to optimize HF management.

**Sources of Funding:** This study was supported by a Grant-in-Aid for Young Scientists [Japan Society for the Promotion of Science KAKENHI, #18K15860 (Y.S.), #23K15168 (Y.S.)], a Grant-in-Aid for Scientific Research © [#23591062 (T.Y.), #26461088 (T.Y.), #16K09469 (Y.N.), #16H05215 (S.K.), #17K09526 (T.K.), #18K08056 (T.Y.), #20K08408 (T.K.), #20H03915 (S.K.)], a Health Labour Sciences Research Grant [#14528506 (S.K.)], the Sakakibara Clinical Research Grant for the Promotion of Sciences [T.Y. 2012-2020], the Japan Agency for Medical Research and Development [201439013C (S.K.)], and the Grant-in-Aid for Clinical Research from the Japanese Circulation Society [Y.S. 2019].

**Disclosures:** The authors declare no conflicts of interest.

## Data Availability

WET-HF registry

https://jcvsd.org/PCI/Login.html

## Acknowledgments

None

## Institutional Review Board Statement

The study protocol was approved by the institutional review boards at each site, and research was conducted in accordance with the Declaration of Helsinki. Ethical approval for the study was granted by the institutional review boards of Keio University Hospital (20090176), Kyorin University (605), Saitama Medical University (15-071), Sakakibara Heart Institute (11-021), St. Luke’s International Hospital (17-R074), Saiseikai Central Hospital (28-55), and National Hospital Organization Tokyo Medical Center (R20-172).

## Non-standard Abbreviations and Acronyms

BMI: body mass index
GDMT: guideline-directed medical therapy
HF: heart failure
HFrEF: heart failure with reduced ejection fraction
HFmrEF: heart failure with mildly reduced ejection fraction
HFpEF: heart failure with preserved ejection fraction
MRA: mineralocorticoid receptor antagonists
RASi: renin-angiotensin-system inhibitors
snLVEF: supra-normal left ventricular ejection fraction
VHD: valvular heart disease
WET-HF Registry: West Tokyo Heart Failure Registry

